# Older age is associated with sustained detection of SARS-CoV-2 in nasopharyngeal swab samples

**DOI:** 10.1101/2020.05.28.20115378

**Authors:** Takeshi Hattori, Masaru Amishima, Daisuke Morinaga, Keisuke Kamada, Sho Nakakubo, Yu Yamashita, Yasuo Shichinohe, Shinichi Fujisawa, Mutsumi Nishida, Yasuyuki Nasuhara, Takanori Teshima, Satoshi Konno

## Abstract

PCR testing of nasopharyngeal swab samples is used for the diagnosis of coronavirus disease 2019 (COVID-19) and for determining timing of discharge. The viral load usually declines at convalescent phase, but sometimes remained positive for a long time even after relief of symptoms. In this study, we identified older age is associated with sustained detection of SARS-CoV-2 in nasopharyngeal swab samples.

## Introduction

Currently, the standard for diagnosis of Severe Acute Respiratory Syndrome-Coronavirus-2 (SARS-CoV-2) infection is a positive result on a polymerase chain reaction (PCR)-based test for virus nucleic acids performed on nasopharyngeal swab samples. PCR test is also used as a guide for patient discharge from designated hospitals and medical institutions. We report here the case of a 97-year-old female who developed mild symptoms and was diagnosed with coronavirus disease 2019 (COVID-19). Although her clinical symptoms and radiological findings resolved within a few days, PCR results from nasopharyngeal swab samples remained positive for 40 days after the onset. This case prompted us to conduct a retrospective study of the association of age the duration of positive PCR testing of nasopharyngeal swab samples in 66 patients diagnosed with COVID-19. We hypothesized that old age could be a risk for prolonged duration of positive PCR results from nasopharyngeal swab samples.

## Methods

This study was approved by the ethics committees of National Hospital Organization Hokkaido Medical Center.

### Sample collection

Nasopharyngeal swab samples were collected from all patients; additionally, oral mucosal swab samples were collected from one patient (the case presentation). Samples were obtained by using FLOQSwabs (COPAN, Murrieta, California, United States [US]). To perform sampling, the swab was passed through the nostril until reaching to the posterior nasopharynx; the swab was removed slowly while rotating. The swabs were placed in saline solution prior to evaluation.

### PCR

Quantitative real-time reverse transcription–PCR (RT-qPCR) was conducted as instructed in the manual for the Detection of Pathogen 2019-nCoV Ver.2.9.1, March 19, 2020, by the National Institute of Infectious Diseases in Japan (https://www.niid.go.jp/niid/images/lab-manual/2019-nCoV20200319.pdf, accessed on 2020-5-3). Total RNA was extracted by QIAamp Viral RNA Mini Kit (QIAGEN, Hilden, Germany) from each specimen. One-step RT-qPCR was performed using One-Step Real-Time RT-PCR Master Mixes (Thermo Fisher Scientific, Waltham, Massachusetts, US) and the extracted RNA. The instrument used was the StepOnePlus Real-time PCR System (Thermo Fisher Scientific). A forward primer (5-AAATTTTGGGGACCAGGAAC-3), reverse primer (5-TGGCAGCTGTGTAGGTCAAC-3) and TaqMan probe (5’-FAM-ATGTCGCGCATTGGCATGGA-BHQ-3’) were used for detection of viral RNA. In a real-time PCR assay, cycle threshold (Ct) value is defined as the number of cycles required for the fluorescent signal to cross a baseline threshold. The test results of SARS-CoV-2 were reported as negative in tests in which Ct>45).

### Patients

The patient in the case presentation has been followed in Hokkaido University Hospital. The patients enrolled in the retrospective study, which explored the relationship between age and duration of time until a positive PCR test reverted to negative, were followed in National Hospital Organization Hokkaido Medical Center. In this hospital, when respiratory physicians (T.H. and M.A.) recognized the peak-out of the COVID-19 infection, PCR tests were performed every day on nasopharyngeal swab samples from each patient. This enabled us to determine the precise day that each COVID-19 patient tested “negative” by PCR. We defined “negative PCR” as confirmed negative results over two sequential days.

### Statistical analysis

Continuous data were presented as means and standard deviations (SD) or medians and interquartile ranges (IQR), while categorical data were presented as frequencies and proportions. To compare differences among the groups, Student’s *t*-tests or one-way analysis of variance were used for parametric continuous variables, Mann–Whitney U tests were used for nonparametric continuous variables, and chi-square tests were used for categorical variables. To evaluate the correlation between age and the duration of positive PCR results, the Pearson’s correlation coefficient was used. To adjust the effect of severity of the disease and the medication used, multivariate regression analysis was applied Statistical analyses were performed using the statistical software package JMP version 13 (SAS Institute Inc., Cary, NC) and SAS version 9.3 (SAS institute, Cary, NC, USA). For all statistical analyses, P < 0.05 was considered significant.

## Case presentation

A 97-year-old woman with a high fever (38□) and diminished appetite for one week was diagnosed with COVID-19 by a positive nasopharyngeal swab PCR test and was referred to Hokkaido University Hospital for ongoing evaluation and treatment. At the time of admission, she was under treatment for hypertension and heart failure. She had no history of smoking. On admission, her blood pressure was slightly high (157/75 mmHg) but she had no cough or dyspnea, and her oxygenation was within normal limits. Laboratory data showed slight elevation of serum C-reactive protein (0.49 mg/dL). Her chest radiograph and computed tomography (CT) imaging revealed slight ground glass opacity (GGO) in the lower left lobe of the lung. In our hospital, we confirmed the positive PCR for SARS-CoV-2 in samples taken from nasopharyngeal swab. Treatment with favipiravir and ciclesonide was initiated; fever resolved within a few days after admission. On hospital day 5, serum CRP decreased to within normal range and the GGOs on chest radiograph disappeared. Her appetite also recovered and here general condition was good. However, at 40 days after the onset of disease, results of PCR on the nasopharyngeal swab samples remain positive. We also confirmed that, results of PCR on the oral swab sample was negative at 35 days. Table 1 documents the change in Ct value over time for PCR tests performed on both oral mucosa and nasopharyngeal swab samples.

**Table 1.** Serial changes in the Ct value over time for PCR tests performed on both oral mucosa and nasopharyngeal swab samples.

| Day after onset | 13 | 15 | 18 | 20 | 23 | 26 | 29 | 35 | 41 |
| --- | --- | --- | --- | --- | --- | --- | --- | --- | --- |
| nasopharyngeal swab sample. | 27.2 | 27.8 | 32.9 | 29.4 | 34.8 | 31.0 | 30.6 | 36.6 | 33.2 |
| oral mucosa swab sample |  |  |  | 35.3 |  |  |  | >45 |  |

The test results of SARS-CoV-2 were reported as negative in tests in which Ct >45).

## Retrospective evaluation of the association between age and the duration of positive PCR tests of samples from nasopharyngeal swabs

The aforementioned case prompted us to conduct a retrospective study to determine the association of patient age with prolonged duration of positive PCR results among patients diagnosed with COVID-19. We analyzed the records of 66 patients who were diagnosed with COVID-19 between March 1, 2020 and April 30, 2020 at National Hospital Organization, Hokkaido Medical Center.

The median age of COVID-19 patients was 61 years old (range, 22 to 99 years old). All patients were admitted to our hospital after having been diagnosed by positive PCR results from nasopharyngeal samples. 42 subjects were mild cases, who did not require supplemental oxygen treatment. 18 subjects were moderate cases who needed oxygen treatment. 6 subjects were severe cases who needed ventilator or/and ECMO. Characteristics of the 66 patients diagnosed with mild COVID-19 are presented in Table 2. Twenty-three subjects had received drug treatment for COVID-19, including favipiravir, litoravir/rapoinavir, ciclesonide, and camostat mesylate.

**Table 1.** Clinical characteristic of subjects diagnosed as COVID-19 in National Hospital Organization Hokkaido Medical Center.

|  | Total (n=66) | Mild (n=42) | Moderate (n=18) | Severe (n=6) | p value* |
| --- | --- | --- | --- | --- | --- |
| Male, n(%) | 38 (57.6) | 23 (54.8) | 12 (66.7) | 3 (7.9) | 0.6421 |
| Age | 61 (22-99) | 56 (28-99) | 71 (30-96) | 59 (22-61) | 0.0553 |
| LDH | 260.5 (127-555) | 249.5 (127-452) | 330.5 (165-555) | 314 (192-452) | 0.2927 |
| D-dimer | 0.75 (0.05-20.3) | 0.6 (0.05-5) | 1.8 (0.4-11.3) | 3.25 (0.2-20.3) | 0.0076 |
| CRP | 3.495 (0.02-25.82) | 2.1 (0.02-17.68) | 5.82 (0.28-24.49) | 5.54 (1.02-25.82) | 0.0122 |
| Ferritin | 293.3<br>(12.5-1760.3) | 268.9<br>(12.5-1760.3) | 402.9<br>(75.1-156.9) | 606.3<br>(225.5-837.9) | 0.1452 |
| HbA1c | 5.9 (5.1-11.1) | 5.9 (5.1-11.1) | 6.15 (5.4-8.0) | 6.85 (5.7-9.9) | 0.173 |
| WBC | 4.9 (1.9-14.8) | 4.8 (1.9-9.1) | 4.8 (2.6-11.6) | 6.95 (3.5-14.8) | 0.2178 |
| bas | 0.2 (0-3) | 0.2 (0-3) | 0.15 (0-0.5) | 0.2 (0-0.4) | 0.7529 |
| eos | 0.05 (0-4.5) | 0.05 (0-3.9) | 0.1 (0-2.1) | 0.1 (0-4.5) | 0.9102 |
| lym | 20.85 (5.7-58.0) | 24.7 (8.8-58) | 15.4 (6-27) | 11.2 (5.7-20.7) | 0.0009 |
| mon | 6.1 (1.5-15.0) | 6.4 (2-11.9) | 5.4 (1.5-15) | 6.05 (1.9-8) | 0.2372 |
| neu | 71.8 (34.0-92.5) | 68.65 (34-88) | 76.85 (60-92.5) | 80.7 (68.8-92) | 0.0028 |
| Medication (for COVID-19) | 36 (54.5) | 17 (40.5) | 14 (77.8) | 5 (54.6) | 0.0097 |
| Ciclesonide | 32 (48.5) | 16 (38.1) | 12 (66.7) | 4 (66.7) | 0.0824 |
| Favipiravir | 21 (31.8) | 5 (11.9) | 12 (66.7) | 4 (66.7) | <0.0001 |
| Ritonavir/Lopiravir | 8 (12.1) | 2 (4.8) | 2 (11.1) | 4 (66.7) | <0.0001 |
| Camostat | 3 (4.6) | 1 (2.4) | 2 (11.1) | 0 (0) | 0.2826 |
| Prednisolone | 2 (3.0) | 0 (0) | 2 (11.1) | 0 (0) | 0.0639 |
| Duration of positive PCR | 19 (9-45) | 17.5 (9-29) | 21 (14-45) | 24.5 (15-43) | 0.0043 |

\* Comparisons among three groups (mild, moderate, and severe)

We found that older age was significantly associated with prolonged positive PCR tests among those diagnosed with COVID-19 (P = 0.0053; Figure 1A). This relationship remained unchanged when the findings were adjusted for the potential impact of severity of the disease (mild, moderate, severe) and the used of medication (P = 0.026). When we analyzed only mild cases of COVID-19 in order to exclude the influence of disease severity, the result remained significants (P = 0.036; Figure 1B).

**Figure 1.**
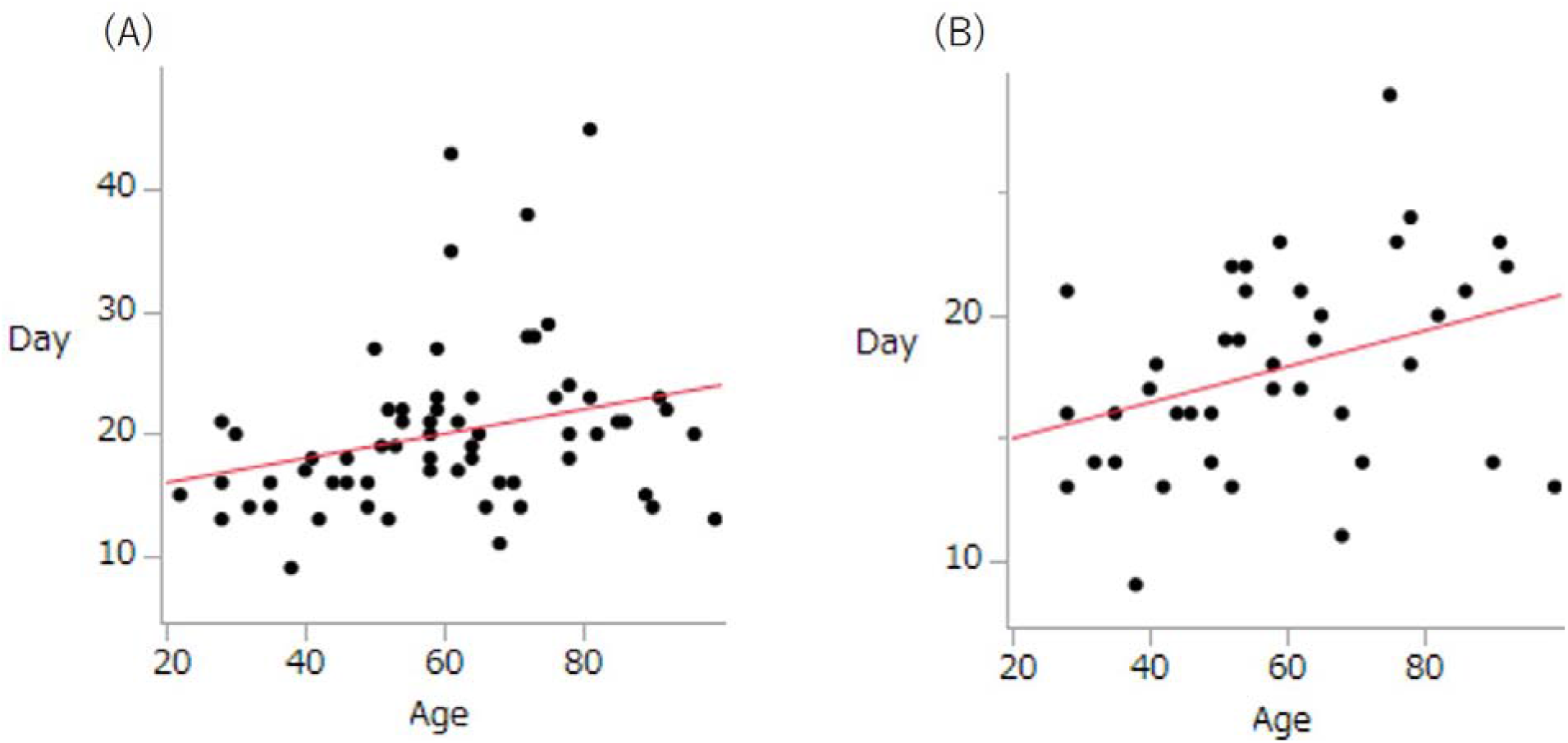
The association of patient age with prolonged duration of positive PCR results among patients diagnosed with COVID-19 at National Hospital Organization, Hokkaido Medical Center. (A) All cases (N=66) (B) Mild cases (N=42)

## Discussion

At the time of this writing, Hokkaido, most northern island in Japan, has controlled the second wave of COVID-19 and the rate of new cases continues to decrease. However, as per the statements of and guidance from the World Health Organization (WHO) (1), patients are discharged only upon confirmed negative PCR tests of nasopharyngeal swabs taken on two sequential days. This results in prolonged hospital stays as patients remain in beds in the designated infectious disease units designed for acute and ongoing care. We have found that quite a few patients are in good general condition, but remain hospitalized due to prolonged positive PCR tests. These patients who no longer need acute care but cannot be discharged are currently a tremendous burden for our hospital.

In our analysis, older age is significantly associated with prolonged duration of positive PCR tests from nasopharyngeal swab samples, irrespective of the disease severity and the medication used (Figure 1). The reasons underlying these observations remain unclear. Of note, we recently compared SARS-CoV-2 detection in nasopharyngeal *vs*. saliva samples from patients diagnosed with COVID-19 (2); we found that quality of PCR from saliva as a diagnostic measure of SARS-CoV-2 infection was equivalent to that of the samples from nasopharyngeal swabs. We also found that findings from PCR tests reverted from positive to negative much more quickly when using saliva than nasopharyngeal samples (2). We speculate that in older individuals, cell turnover is less robust and as such, clearance of virus from the nasopharynx is prolonged; these factors may lead to positive PCR tests that persist after acute disease has resolved. We note that the PCR test on an oral mucosal swab sample was negative on hospital day 35. It is also possible that PCR detects nonviable particles of viral components in less clean environment such as nasopharynx compared to oral cavity.

One group from Taiwan has already discussed the possibility that COVID-19 may no longer be contagious at two weeks after the onset of symptoms (3). As such, it is possible the positive PCR tests reflect the presence of that inactive virions remaining within nose. Their findings suggest that most transmission of COVID-19 occurred at the very early stages of the disease and perhaps even before the onset of symptoms; of note, the secondary clinical attack rate among contacts decreased over time as symptoms developed and progressed. In a linked study, Zheng and colleagues evaluated viral loads in respiratory samples, stool, serum, and urine using PCR: they found that more than half the respiratory samples remained positive for SARS-CoV-2 as did a full one third of the stool samples at the end of the four week trial period (4). One important limitation of PCR testing is the inability to differentiate between active infectious virus and their non-viable, non-infectious viral counterparts. As such, we propose that there should be a change in the strategy currently in use for determining time of discharge to one that relies on other clinical tests or/and patients’ condition, which could be helpful to inhibit the development of patients’ flail and dementia and to reduce the burden of ongoing and potentially unnecessary prolonged hospitalization.

In summary, it became evident that the results from nasopharyngeal swab PCR tests remained positive for SARS-CoV-2 for prolonged periods of time despite resolution of the associated clinical illness. Further, we demonstrated that old age is significantly associated with prolonged duration of positive PCR results from nasopharyngeal swab samples; this is the case regardless of disease severity. Further studies will be needed in order to clarify how long these patients are actually contagious; this will be critical toward the effort to relieve the comparatively large burden placed on hospitals and hospital staff at this time.

## Data Availability

No available data was included.

## References

1 World Health Organization. Home care for patients with COVID-19 presenting with mild symptoms and management of their contacts. Interim guidance 17 March 2020.

2 Comparison of SARS-CoV-2 detection in nasopharyngeal swab and saliva. Iwasaki S, Fujisawa S, Nakakubo S, Kamada K, Yamashita Y, Fukumoto T, Sato K, Ogur S, Taki K, Senjo H, Sugita J, Hayasaka K, Konno S, Nishida M, Teshima T. medRxiv preprint. doi:https://doi.org/10.1101/2020.05.13.20100206.

3 Cheng HY, Jian SW, Liu DP, Ng TC, Huang WT, Lin, MD. Contact tracing assessment of COVID-19 transmission dynamics in Taiwan and risk at different exposure periods. JAMA Int Med. 2020 1.

4 Roos E, Marieke JA. Persistence of viral RNA in stool samples from patients recovering from covid-19 PCR has limitations, and isolating patients for a month or more may not be feasible isolation. BMJ 369 m1724.

